# Covaxin (BBV152) Vaccine Neutralizes SARS-CoV-2 Delta and Omicron variants

**DOI:** 10.1101/2022.01.24.22269189

**Authors:** Venkata-Viswanadh Edara, Mit Patel, Mehul S. Suthar

## Abstract

The SARS-CoV-2 vaccine BBV152/Covaxin is well-tolerated and was shown to be 77.8% efficacious against symptomatic and 93.4% efficacious against severe symptomatic COVID-19 disease in adults. Previous studies have shown that sera from Covaxin vaccinated individuals have neutralizing activity against B.1.1.7 (Alpha), B.1.351 (Beta), B.1.617.2 (Delta), B.1.1.28 (Zeta), and B.1.617.1 (Kappa) SARS-CoV-2 variants. The B.1.1.529 variant (Omicron) recently emerged in November 2021 and has spread throughout the world. The Omicron variant has more than 30 mutations within the spike protein that could impact vaccine-mediated immunity. We used a live virus neutralization assay to evaluate the neutralizing activity against the Omicron variant of sera collected from subjects who received a booster dose (6-month after primary series last dose) of Covaxin. We found that sera from Covaxin boosted individuals showed neutralizing activity against D614G (vaccine strain), Delta, and Omicron variants. One hundred percent of boosted subjects showed neutralizing activity against the Delta variant while over 90% of boosted subjects showed neutralizing activity against the Omicron variant. These findings show that a booster dose of Covaxin can generate robust neutralizing antibody responses against the Omicron variant.

## Main

The SARS-CoV-2 vaccine BBV152/Covaxin is well-tolerated and was shown to be 77.8% efficacious against symptomatic and 93.4% efficacious against severe symptomatic COVID-19 disease in adults.^1^ SARS-CoV-2 variants with mutations throughout the genome, including the spike protein, have emerged and spread throughout the world. The Alpha (B.1.1.7), Beta (B.1.351), Gamma (B.1.1.28), Delta (B.1.617.2), and Omicron (B.1.1.529) variants pose serious public health concerns due to higher transmissibility, immune escape, and disease severity. The Omicron variant has more than 30 mutations within the spike protein. There is increase in breakthrough infection in people vaccinated with primary series dosing regimen and those who received booster doses.^2,3^ Reduced neutralizing activity of antibodies against Omicron variant may potentially lead to symptomatic infection, increased disease severity and hospitalization.

BBV152/Covaxin is an inactivated whole virion vaccine that induces robust antibodies and cell-mediated immune memory responses. Earlier studies have shown neutralization potential of the Covaxin vaccinee’s sera against Alpha (B.1.1.7), Beta (B.1.351), Delta (B.1.617.2), Zeta (B.1.1.28), and Kappa (B.1.617.1) variants of SARS-CoV-2.^5-8^ Here we used a live virus focus reduction neutralization test (FRNT) to evaluate the neutralizing activity of sera collected from subjects who received a booster dose (6-month after primary series last dose) of Covaxin against the Omicron variant.

In an ongoing phase 2 trial (ClinicalTrials.gov: NCT04471519) the protocol was amended after six months to re-consent and randomize previously vaccinated participants to receive a third dose (booster) of Covaxin on Day 215.^9^ Sera were collected 28 days after booster and used in neutralization assay in this study. Sera from booster study was effective in neutralizing the D614G (vaccine strain), Delta, and Omicron variants. In the boosted individuals that received Covaxin, the Geometric Mean Titers (GMTs) were 706 for D614G, 480 for Delta and 75 for Omicron and corresponded to a 1.5- and 9.4-fold reduction as compared to D614G, respectively (Figure 1). One hundred percent of boosted subjects showed neutralizing activity against the Delta variant, while over 90% of boosted subjects showed neutralizing activity against the Omicron variant.

**Figure 1.**
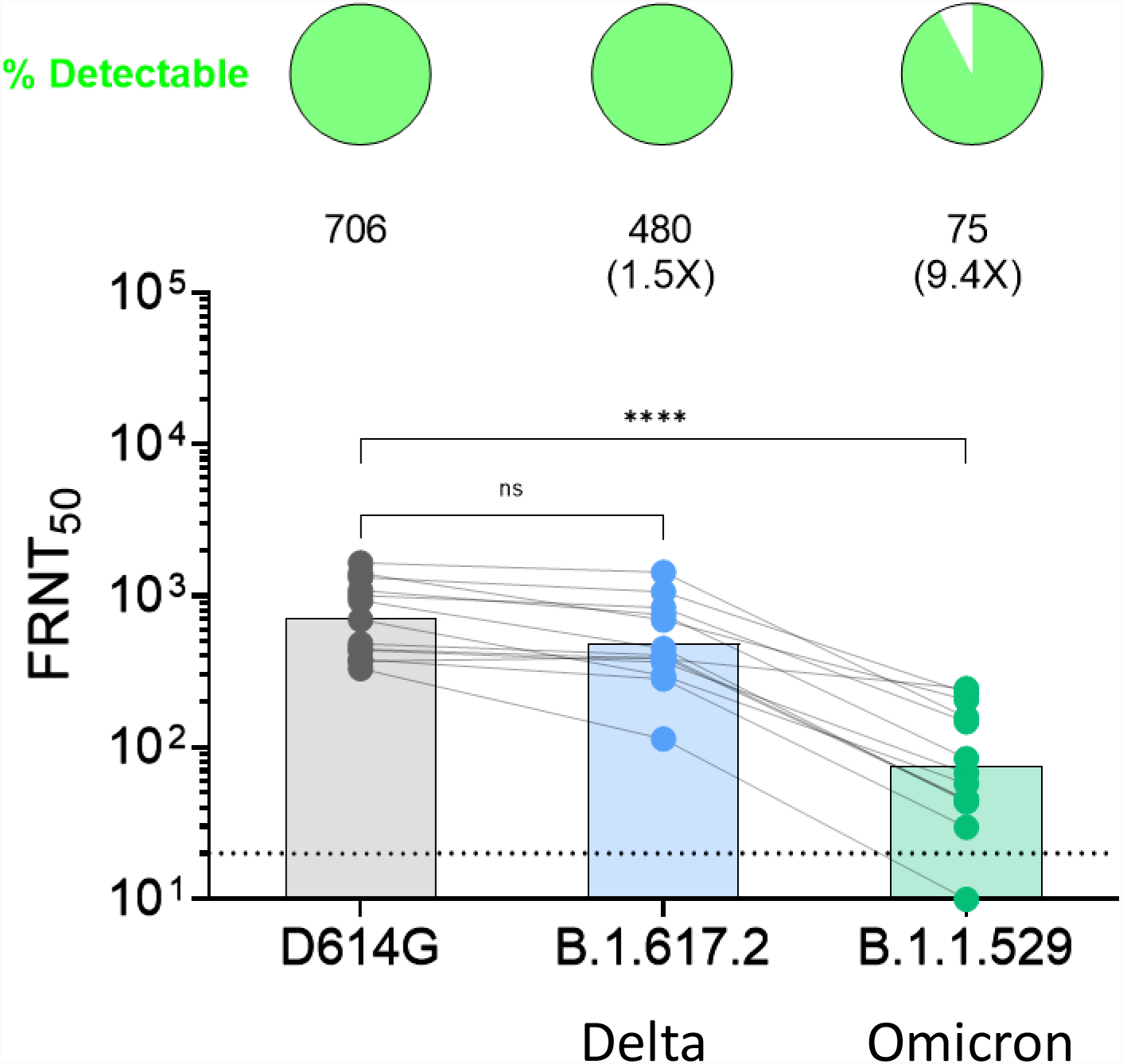
Neutralization antibody responses against D614G, B.1.617.2 (Delta), and B.1.1.529 (Omicron) SARS-CoV-2 variants post-Covaxin booster dosing. Sera samples (28 days after booster dose) collected from Covaxin vaccinated individuals that received a booster dose (n=13) at 6-months following 2-dose primary series. The FRNT50 GMTs for D614G, Delta, and Omicron are shown with respective fold changes in comparison to D614G strain. A pie-chart above each graph shows the frequency of individuals that have detectable titers above the limit of detection (LOD). The connecting lines between the variants represents matched serum samples. The horizontal dashed lines along the X-axis indicate the limit of detection (FRNT50 GMT= 10). Non-parametric pairwise analysis for neutralization titers were performed by Kruskal-Wallis test with Dunn’s multiple comparisons test. ****p<0.0001

These findings show that a booster dose of Covaxin can generate neutralizing antibody responses against the Omicron variant. Sera from booster study were effective in neutralizing the Delta and Omicron variants. The neutralization activity of Covaxin boosted sera against Omicron was comparable to mRNA vaccine boosted sera using the same live virus neutralization assay.^10^ This study highlights the importance of booster dosing in improving waning neutralizing antibodies in individuals that have received the primary vaccine series.^9^

The limitations of this study include: 1) small sample size; 2) we are not able to link the clinical outcomes with the neutralization findings; 3) this study did not evaluate T cell immunity which likely plays an important role in protection against COVID-19.

## Data Availability

All data produced in the present study are available upon reasonable request to the authors

## Acknowledgements

This work was sponsored by Ocugen, Inc., Malvern, USA, and serum samples were provided by Bharat Biotech India Limited, Hyderabad, India.

## Methods

### Viruses and cells

VeroE6-TMPRSS2 cells were generated and cultured as previously described.^11^ VeroE6-TMPRSS2 cells were used to propagate all virus stocks. The D614G, B.1.617.2, and B.1.1.529 viruses were isolated and propagated on Vero-TMPRSS2 cells as previously described.^10-12^ All the variants were plaque purified directly from the nasal swabs, propagated once in a 12-well plate, and expanded in a confluent T175 flasks to generate working stocks. All viruses used in this study were deep sequenced and confirmed as previously described.^11^

### Samples

Collection and processing were performed by Bharat Biotech India Limited. These samples were collected from booster study. In the ongoing phase 2 trial (ClinicalTrials.gov: NCT04471519) the protocol was amended after six months to re-consent and randomize previously vaccinated participants to receive a third dose (booster) of Covaxin on Day 215. Sera were collected 28 days after booster and used in neutralization assay in this study.^9^

### Focus Reduction Neutralization Test

FRNT assays were performed as previously described.^11,13,14^ Briefly, samples were diluted at 3-fold in 8 serial dilutions using DMEM in duplicates with an initial dilution of 1:10 in a total volume of 60 μL. Serially diluted samples were incubated with an equal volume of D614G, B.1.617.2, and B.1.1.529 (100-200 foci per well based on the target cell) at 37°C for 45 minutes in a 96-well plates. The antibody-virus mixture was added to VeroE6-TMPRSS2 cells and incubated at 37°C for 1 hour. Post-incubation, the antibody-virus mixture was removed and pre-warmed 0.85% methylcellulose overlay was added to each well. Plates were incubated at 37°C at 18 hours for D614G and Delta variants and 40 hours for the Omicron variant. At this time, the methylcellulose overlay was removed and washed six times with PBS. Cells were fixed with 2% paraformaldehyde in PBS for 30 minutes. Following fixation, plates were washed twice with PBS and permeabilization buffer (0.1% BSA, Saponin in PBS) was added to permeabilized cells for at least 20 minutes. Cells were incubated with an anti-SARS-CoV spike primary antibody directly conjugated to Alexa-flour-647 (CR3022-AF647) overnight at 4°C. Cells were washed three times in PBS and foci were visualized on a CTL Analyzer.

### Quantification and Statistical Analysis

Antibody neutralization was quantified by counting the number of foci for each sample using the Viridot.^15^ The neutralization titers were calculated as follows: 1 - (ratio of the mean number of foci in the presence of sera and foci at the highest dilution of respective sera sample). Each specimen was tested in duplicate. The FRNT50 titers were interpolated using a 4-parameter nonlinear regression in GraphPad Prism 9.2.0. Samples that do not neutralize at the limit of detection at 50% are plotted at 10 and were used for geometric mean and fold-change calculations.

## References

1. Ella, R., et al. Efficacy, safety, and lot-to-lot immunogenicity of an inactivated SARS-CoV-2 vaccine (BBV152): interim results of a randomised, double-blind, controlled, phase 3 trial. Lancet 398, 2173–2184 (2021).

2. Wang, S.Y., et al. Severe breakthrough COVID-19 cases in the SARS-CoV-2 delta (B.1.617.2) variant era. Lancet Microbe 3, e4–e5 (2022).

3. Goldberg, Y., et al. Protection and waning of natural and hybrid COVID-19 immunity. medRxiv, 2021.2012.2004.21267114 (2021).

4. CDC. COVID-19 Vaccines Work. Vol. 2022 (Centers for Disease Control and Prevention, 2021).

5. Sapkal, G.N., et al. Inactivated COVID-19 vaccine BBV152/COVAXIN effectively neutralizes recently emerged B.1.1.7 variant of SARS-CoV-2. J Travel Med 28(2021).

6. Sapkal, G., et al. Neutralization of VUI B.1.1.28 P2 variant with sera of COVID-19 recovered cases and recipients of Covaxin an inactivated COVID-19 vaccine. J Travel Med 28(2021).

7. Yadav, P.D., et al. Neutralization of variant under investigation B.1.617 with sera of BBV152 vaccinees. Clin Infect Dis (2021).

8. Yadav, P.D., et al. Neutralization of Beta and Delta variant with sera of COVID-19 recovered cases and vaccinees of inactivated COVID-19 vaccine BBV152/Covaxin. J Travel Med 28(2021).

9. Vadrevu, K.M., et al. Persistence of immunity and impact of a third (booster) dose of an inactivated SARS-CoV-2 vaccine, BBV152; a phase 2, double-blind, randomised controlled trial. medRxiv, 2022.2001.2005.22268777 (2022).

10. Edara, V.-V., et al. mRNA-1273 and BNT162b2 mRNA vaccines have reduced neutralizing activity against the SARS-CoV-2 Omicron variant. bioRxiv, 2021.2012.2020.473557 (2021).

11. Edara, V.V., et al. Infection and Vaccine-Induced Neutralizing-Antibody Responses to the SARS-CoV-2 B.1.617 Variants. N Engl J Med 385, 664–666 (2021).

12. Edara, V.V., Hudson, W.H., Xie, X., Ahmed, R. & Suthar, M.S. Neutralizing Antibodies Against SARS-CoV-2 Variants After Infection and Vaccination. JAMA 325, 1896–1898 (2021).

13. Vanderheiden, A., et al. Development of a Rapid Focus Reduction Neutralization Test Assay for Measuring SARS-CoV-2 Neutralizing Antibodies. Curr Protoc Immunol 131, e116 (2020).

14. Edara, V.V., et al. Infection- and vaccine-induced antibody binding and neutralization of the B.1.351 SARS-CoV-2 variant. Cell Host Microbe 29, 516–521 e513 (2021).

15. Katzelnick, L.C., et al. Viridot: An automated virus plaque (immunofocus) counter for the measurement of serological neutralizing responses with application to dengue virus. PLoS Negl Trop Dis 12, e0006862 (2018).

